# Dramatic increases in redundant publications in the Generative AI era

**DOI:** 10.1101/2025.09.09.25335401

**Authors:** Danny Maupin, Tulsi Suchak, Adrian Barnett, Matt Spick

## Abstract

**Background:** Redundant publication, the practice of submitting the same or substantially overlapping manuscripts multiple times, distorts the scientific record and wastes resources. Since 2022, publications using large open-science data resources have increased substantially, raising concerns that Generative AI (GenAI) may be facilitating the production of formulaic, redundant manuscripts. In this work we aim to quantify the extent of redundant publication from a single, large health dataset and to investigate whether GenAI can create submissions that evade standard integrity checks.

**Methods:** We conducted a systematic search for the years 2021 to 2025 (year to end-July) to identify redundant publications using the US Centers for Disease Control and Prevention National Health and Nutrition Examination Survey (NHANES) dataset. Redundancy was defined as publications analysing the same exposures associated with the same outcomes in the same national population. To test whether GenAI could facilitate creating these papers, we prompted large language models to write three synthetic manuscripts using redundant publications from our dataset as input, instructing them to maximise syntactic differences and evade plagiarism detectors. These three synthetic manuscripts were then tested using a leading plagiarism detection platform to assess their similarity scores.

**Findings:** Our search identified 411 redundant publications across 156 unique exposure-outcome pairings; for example, the association between oxidative balance score and chronic kidney disease using NHANES data was published six times in one year. In many instances, redundant articles appeared within the same journals. The three synthetic manuscripts created by GenAI to evade detection yielded overall similarity scores of 26%, 19%, and 14%, with individual similarity contributions below the typical 5% warning thresholds used by plagiarism detectors.

**Interpretation:** The rapid growth in redundant publications (a 17-fold increase between 2022 and 2024) is suggestive of a systemic failure of editorial checks. These papers distort meta-analyses and scientometric studies, waste scarce peer review resources and pose a significant threat to the integrity of the scientific record. We conclude that current checks for redundant publications and plagiarism are no longer fit for purpose in the GenAI era.

## Introduction

Artificial intelligence tools, including both large language models (LLMs) and easily deployable machine learning workflows, offer potential for both new insights and productivity gains to researchers. ^1^ One promising area is large and complex data resources, such as in healthcare and the biosciences. ^2,3^ Inevitably, however, AI will multiply negative as well as positive outputs. ^4^ The publishing industry has always had to contend with unethical behaviours, and guidelines on best practices in publishing are provided by the Committee on Publication Ethics (COPE). These negative behaviours include text and image recycling by authors; ^5^ plagiarism of others’ work; ^6^ redundant submissions, with the latter including salami slicing or “dividing data or research findings into their smallest publishable units to increase the author’s publication output”; ^7,8^ and systematic manipulation. ^9,10^

We and others have previously documented the exploitation of Open Science data resources for the production of misleading research by entities such as paper mills (organizations that produce manuscripts for sale using derivative or fabricated text, images or data), ^11–13^ including via p-hacking and data dredging. ^14,15^ The ability of GenAI to synthesise new text in a handful of clicks poses an additional problem, as it bypasses both text recycling and plagiarism editorial checks, allowing mass manufacture of redundant manuscripts or simultaneous submission to journals. Whilst some publishers have processes to detect duplicate submissions, ^16^ GenAI can assist with evading these checks. Entities such as paper mills might even be able to submit unlimited ***conceptually identical*** but ***syntactically different*** manuscripts, dramatically increasing the potential production rate of redundant publications. When LLMs are combined with large public datasets such as NHANES or the FDA Adverse Events Reporting System (FAERS), the only rate limiting step remaining is the ability of publishers to review and accept manuscripts, placing ever more strain on editors and peer reviewers.

In addition to negative outcomes arising from the use of GenAI by bad actors such as paper mills, there may also be risks from cross-domain work that substitutes traditional human collaboration with AI. LLMs can act as expert summarisers, allowing a researcher in one field to rapidly acquire a functional understanding of another, thereby reducing the need for direct partnership with domain experts. For instance, a researcher looking for evidence to support a hypothesis might use a LLM to write statistical code to interrogate a large open access database. This approach streamlines the preliminary stages of interdisciplinary research, allowing for quick hypothesis generation and project scoping. This represents a clear productivity gain (which will be hard for small teams to pass up in a highly competitive landscape), ^17^ reducing the difficulties associated with assembling and managing cross-disciplinary teams (conflicting terminologies, differing methodological standards, logistical overheads). ^18^ It can, however, also lead to epistemic trespassing, whereby researchers have a surface level understanding but lack deep domain expertise. Whilst this has been previously described for systematic reviews and meta-analyses, ^19,20^ GenAI can enable such opportunistic authorship more broadly.

Here, we aim to investigate whether GenAI is contributing to a breakdown in the effectiveness of integrity checks in the scientific publishing process. First, we examine whether redundant publication is becoming more prevalent, whether this is inadvertent (through epistemic trespassing) or deliberate (through systematic manipulation). We use the NHANES dataset as our primary source ^21^ and investigate whether the incidence of redundant publications from this dataset has changed over time. Second, we investigate whether LLMs can facilitate the duplication of published works, by rewriting existing manuscripts and testing whether they pass plagiarism checks used by major publishers.

## Materials and methods

### Literature search and manuscript matching

PubMed was used as the primary data source, with the following search string “*(nhanes[tiab] OR national health and nutrition examination survey[tiab]) AND ((“2021/01/01”[Date - Publication]* : *“2025/07/31”[Date - Publication]))*”, which produced an initial list of 15 597 publications. Following this, articles referencing the distinct and independent Korean NHANES survey were excluded (n = 1 657). In addition, comments on publications were excluded (n = 50). A list of exposures and outcomes was derived from our previous work on inappropriate manuscripts using the NHANES dataset, ^15^ and this was added to based on the authors’ experience of more recent literature analysing biomarkers and predictors. In total, a list of 217 exposures / predictors and 162 outcomes was generated (Table S1, Supplementary Material), including alternatives where appropriate.

Manuscript titles were searched for matching exposures and predictors by simple token matching, resulting in a set of 3 465 manuscripts analysing the predefined list of simple exposures and outcomes. Those with more than one paper analysing the same exposure and outcome were recorded. Prior to matching, alternatives were manually replaced with general tokens (for example depressive symptoms was converted to depression, as both outcomes are measured using the same NHANES questionnaire). Titles and abstracts were then manually inspected; studies were treated as effectively redundant publications if they analysed the same exposure and outcome with only variations such as cohort included (e.g. 2014–2018 versus 2016–2020), male versus female, or variations in age ranges. More specific investigations, for example measuring associations between an exposure and an outcome in cancer survivors, were not included as redundant publications (n = 3 054), leaving 411 publications which under the definitions used in this work were redundant. This screening process was not intended to identify problematic publications in general (such as salami slicing or plagiarism); this workflow was intended to identify the specific issue of redundant / duplicate publication.

### Investigation of plagiarism detection

In order to pilot whether simple click-science workflows were sufficient to evade existing mainstream plagiarism detection, three ‘new’ manuscripts were generated from existing published papers. The exposure and outcomes chosen are set out in Table 2 alongside the PMIDs of the ‘source’ publications for the synthesis.

**Table 2:** Pairings of exposures and outcomes selected for the production of synthetic manuscripts

| Exposure – outcome pair | LE8 and Periodontitis | BRI and Infertility | Oxidative Balance Score and Chronic Kidney Disease |
| --- | --- | --- | --- |
| Existing number of duplicates | 6 | 5 | 6 |
| PMIDs of existing duplicates | 38964852 |  | 38919480 |
|  | 38681053 | 39545053 | 39830066 |
|  | 38178120 | 39742100 | 39399530 |
|  | 39739130 | 39696159 | 38877058 |
|  | 38622031 | 39827325 | 39752406 |
|  | 39730816 | 39959618 | 38725577 |

*LE8: Life’s Essential 8, BRI: Body Roundness Index, PMID: PubMed unique identifier*

These exposure / outcome pairings were selected as examples due to the high number of duplicates found during pilot searches, and the large amount of available literature on the topics, which represent sources of considerable public interest. First, ‘new’ manuscripts were generated by three of the authors (D.M., T.S. and M.S.) independently using LLMs to rewrite existing manuscripts, with the only requirement that prompts explicitly specify that the new text should be syntactically different to the original text to avoid plagiarism detection. In addition to the main sections, LLMs were used to generate new syntactically different table titles, figure legends and end disclosures. Second, the ‘investigated’ cohort was changed, and all numbers were adjusted either by instructing an LLM to create new tables, or by manual adjustments to reflect the changes that subtle alterations to NHANES cohorts can produce. Third, all text was manually adjusted where appropriate to reflect the numerical changes, to ensure that text referred to the correct figure and table numbers, and to remove repetition of acronyms (for example spelling out LE8 as “Life’s Essential 8 (LE8)” in full each time mentioned) and formatted. It should be noted that the LLMs could not be prompted to produce completely error-free manuscripts, but the overall production process in each case was around two hours per paper.

Subsequently, the three generated documents were submitted to iThenticate to assess whether a major current plagiarism detector would identify them as unoriginal documents. ^22^ We followed guidance from Springer Nature’s documentation on Crossref Similarity Check / iThenticate that it is “more important to look at the individual scores of the sources than the overall similarity index” and that individual scores in the 1% to 5% range offer “in general no sign of potential plagiarism”. ^23^ Therefore, we treated individual scores over 5% as generating potential red flags for editors and reviewers.

## Results

### Literature search and manuscript matching

The literature search and manuscript matching process generated 411 paired redundant exposure-outcome documents. The most common result was two papers covering the same exposure / outcome pairing (n = 190 publications). The largest number of redundant analyses was six, occurring for three pairings (n = 18 publications). The 20 most frequent exposure–outcome pairs and the distribution of redundancies are shown in Figures 3A and 3B respectively. Figure 3C illustrates the incidence of the duplicates identified over time; there were very few redundant publications relating to NHANES prior to 2023 (12 in 2022 and 3 in 2021), with the 2024 count of 198 representing a 17-fold increase in two years.

**Figure 1:**
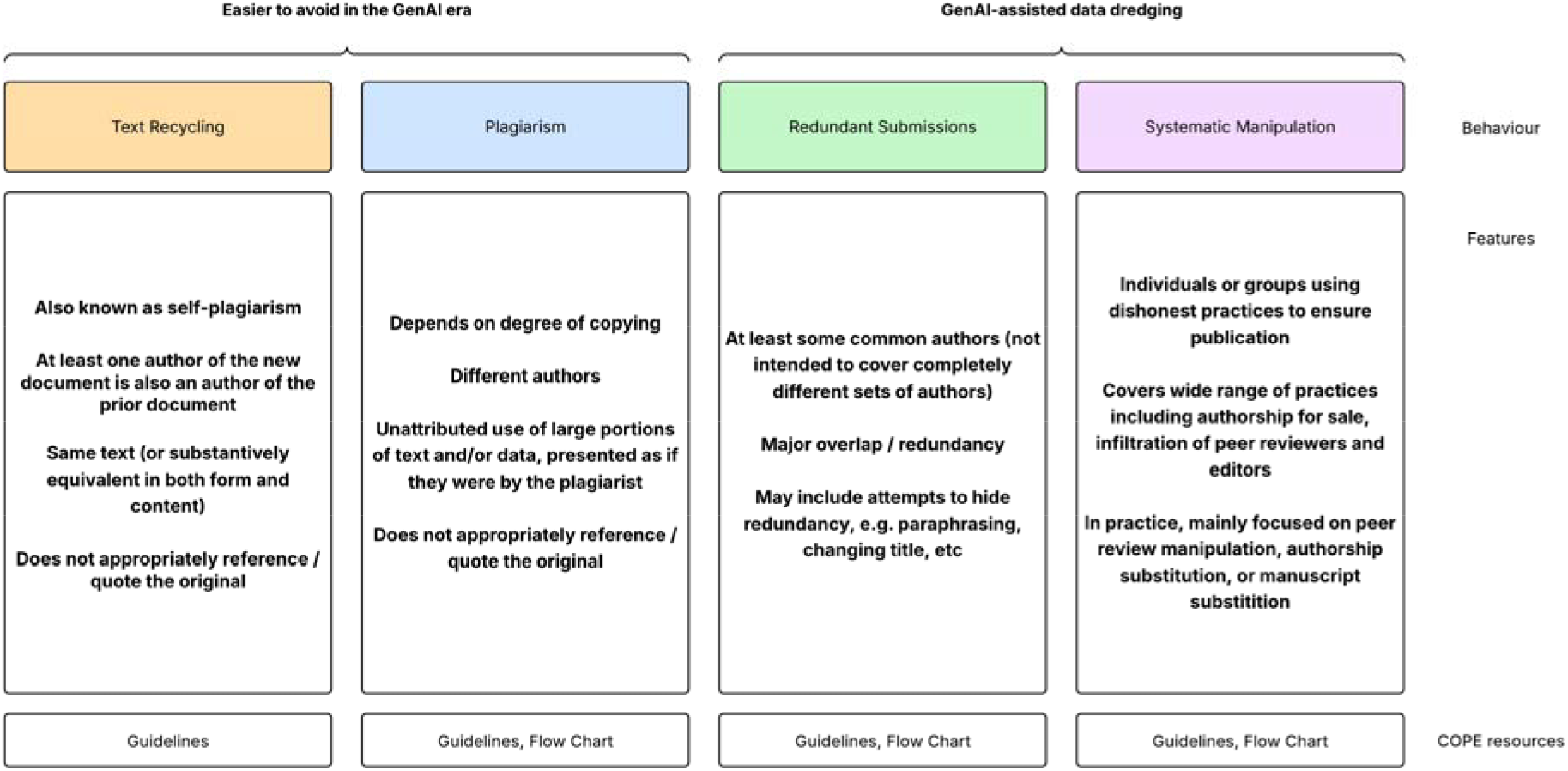
Existing COPE guidelines around plagiarism and related examples of misconduct

**Figure 2:**
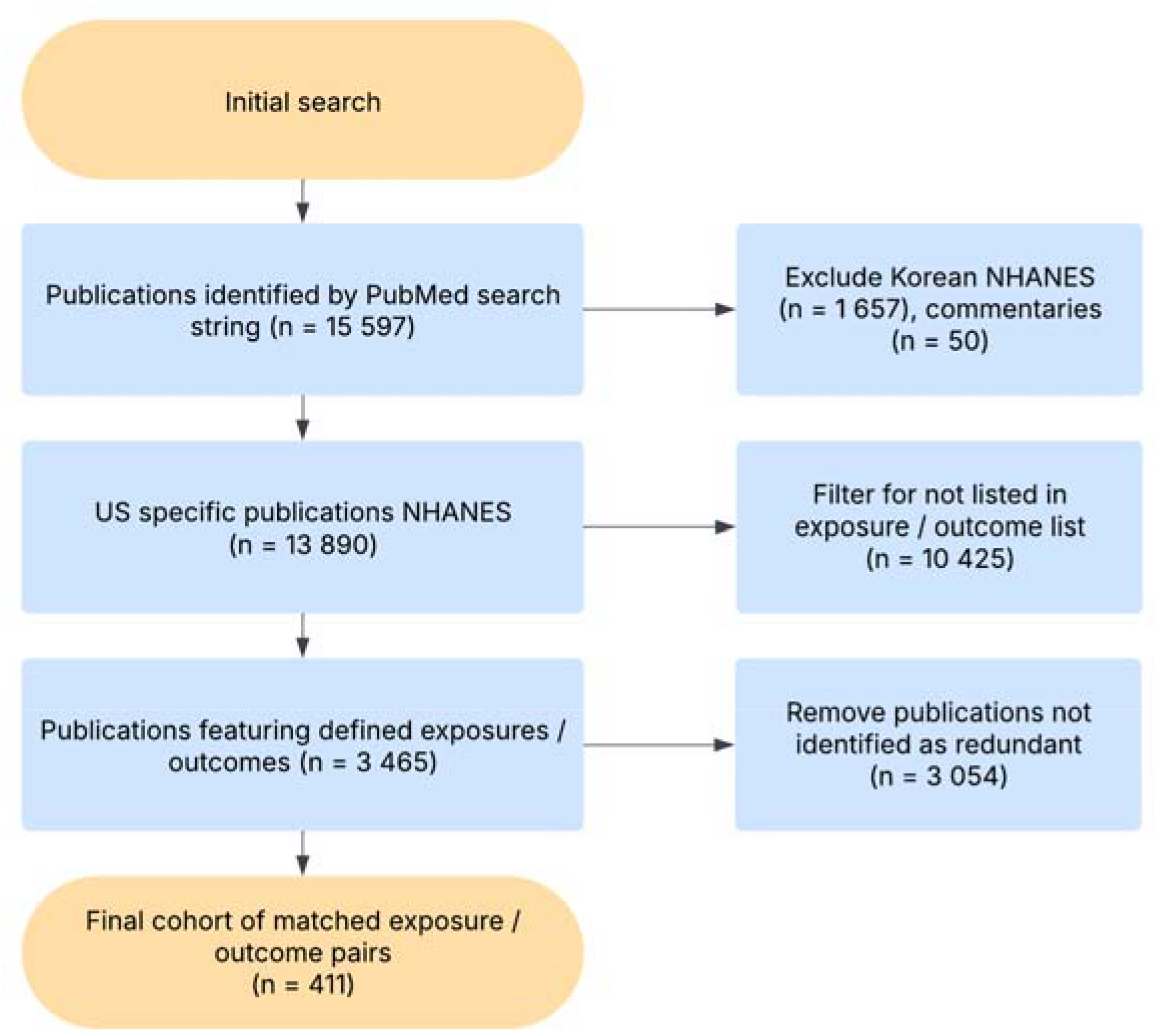
Flowchart of inclusion and exclusion criteria for redundant publication search

**Figure 3:**
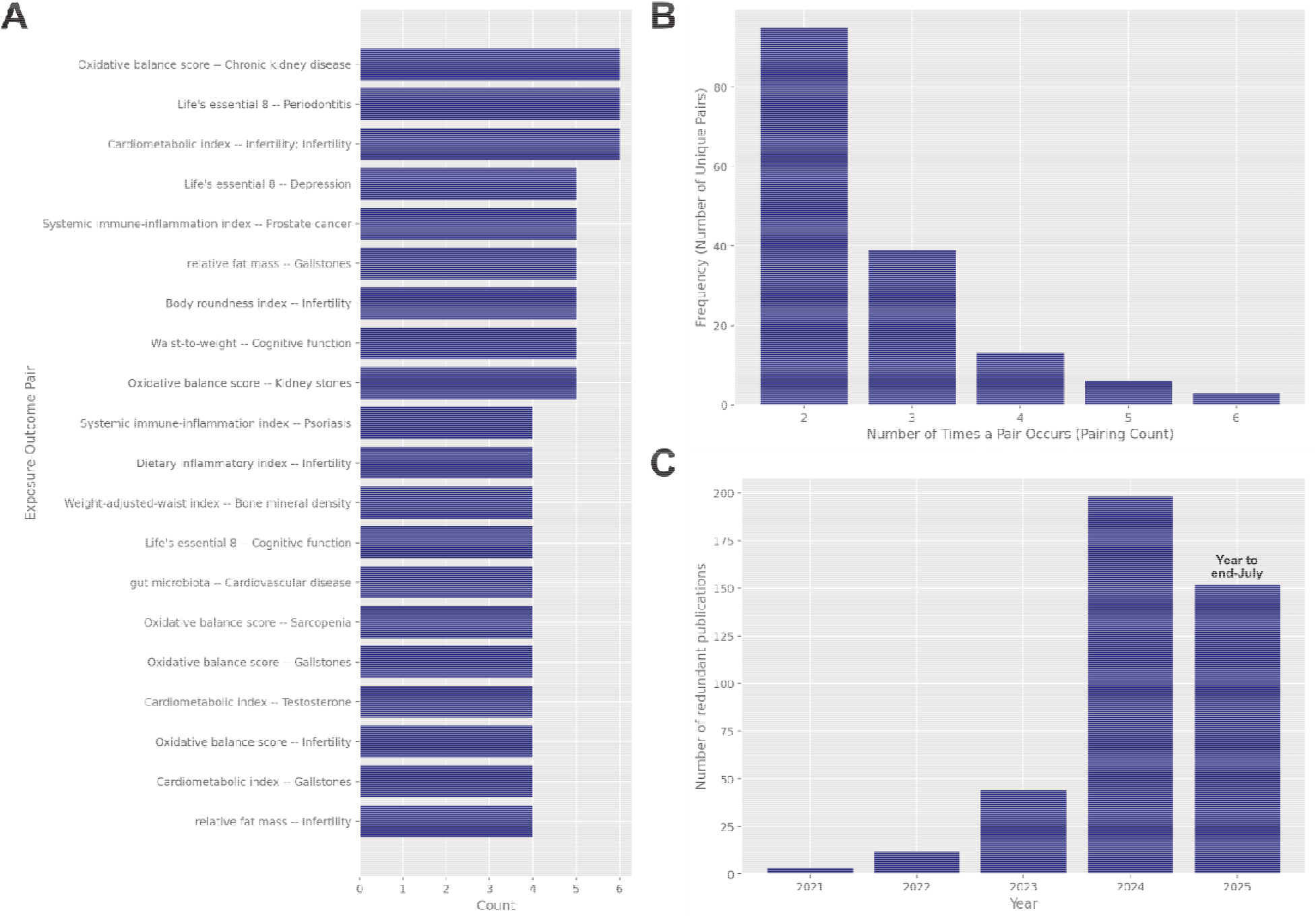
(A) 20 most frequent exposure–outcome pairs (B) Distribution of redundant publications (C) time trend of redundant publications, 2025 is YTD to end-July 2025

The 411 articles were published by 112 journals (Figure 4A), with 44% concentrated in just five journals. The most common statistical method used in the duplicated research publications was logistic regression across multiple models adding covariates cumulatively, followed by linear regression, with a small number of other methods used (Figure 4B).

**Figure 4:**
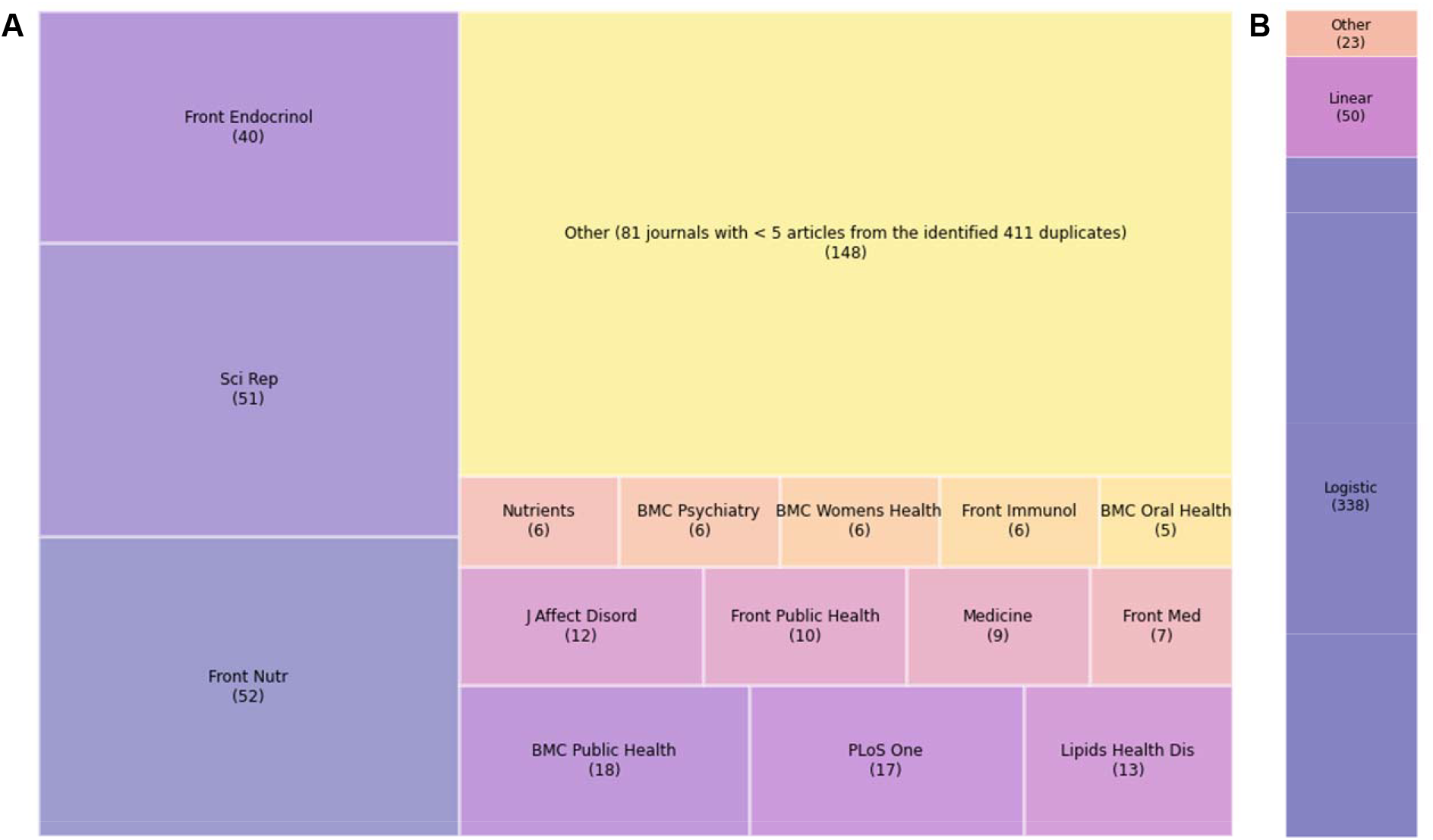
(A) tree map of journals using the number of redundant papers (B) breakdown of statistical methods used in the redundant papers. We use the abbreviated journal name – full journal names are provided in Supplementary Materials, Table S2

### Submission and publication dates

Three illustrative pairings were chosen from those with the highest rates of redundant publications for the generation of synthetic manuscripts. These were chosen as they should be harder to be duplicate and evade plagiarism detection (as more redundant instances are available in the public domain). Submission and publication dates for these redundant publications are shown in Figure 5, together with the journals that published them. Articles commonly followed a pattern of simultaneous or near-simultaneous submission, but also included examples of submissions after the publication of previous works. In some cases, the same journals published the redundant manuscripts. This occurred 67 times across the cohort of 411 publications, with *Frontiers in Nutrition* publishing the same redundant pairing on 29 occasions, and *Scientific Reports* on 10 occasions (the full list of journals is included in Supplementary Materials, Table S2).

**Figure 5:**
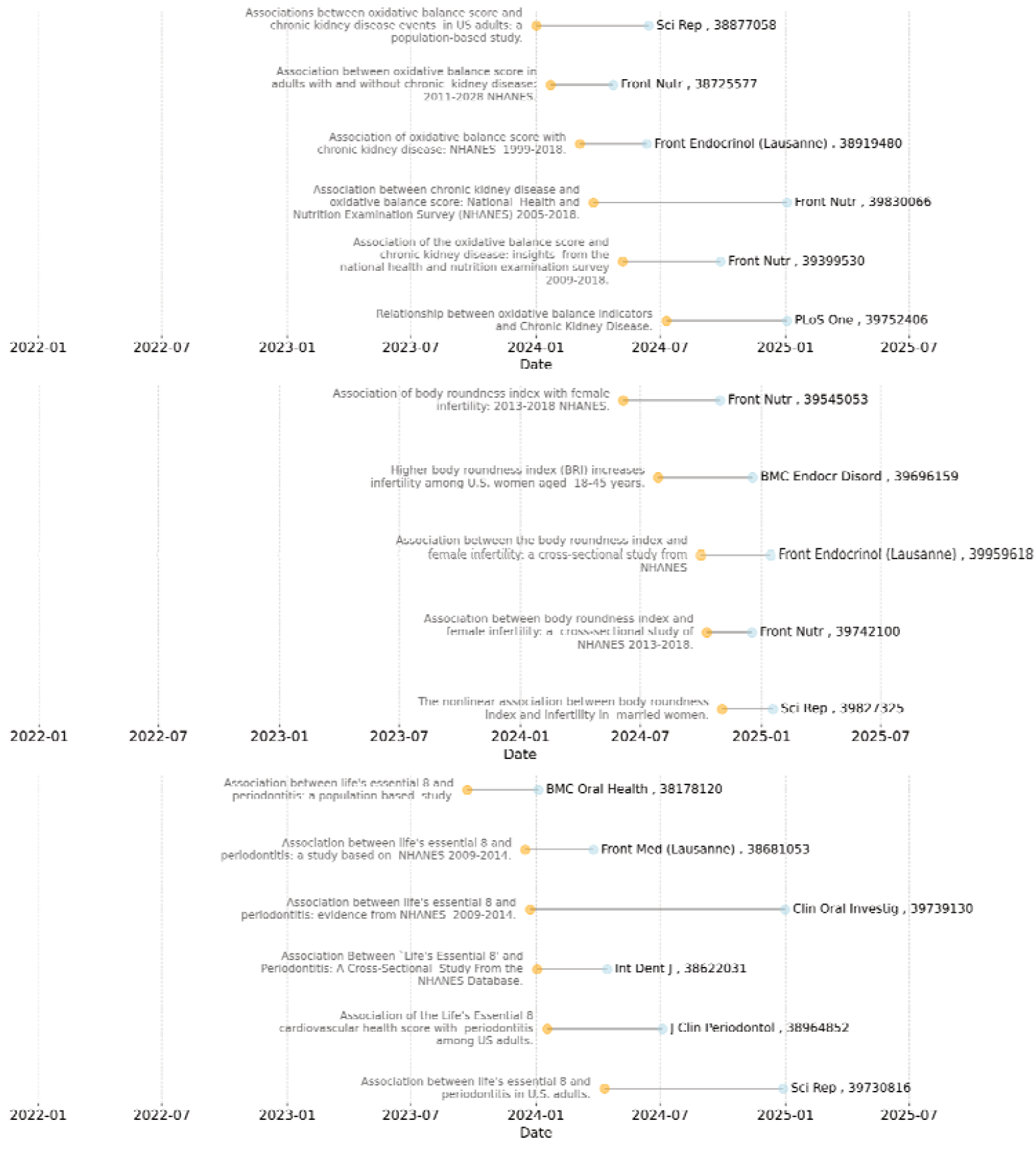
Submission and publication date plots for three outcome–exposure pairings, with publication titles to the left of the submission dates and journal title and publication PMIDs to the right of acceptance dates : (A) oxidative balance score and chronic kidney disease (B) body roundness index and female infertility (C) Life’s Essential 8 and periodontitis

### Evasion of plagiarism checks

The three synthetic manuscripts generated by LLM were then tested using iThenticate. The results are shown in Table 3 for the total similarity scores and for the five main individual similarity sources for each manuscript. The total scores including bibliographical matching are included for reference. All three manuscripts had percentage matching scores that were below 30% overall and no individual source percentages exceeded 5%, and so would not have raised automated red flags in editorial pipelines. ^23^

**Table 3:** Summary of iThenticate checks for three synthetic manuscripts, processed on 26 August 2025

| Exposure – outcome pair | LE8 and Periodontitis | BRI and Infertility | Oxidative Balance Score and Chronic Kidney Disease |
| --- | --- | --- | --- |
| LLM used | ChatGPT 5 | Gemini 2.5 Pro | ChatGPT 4o |
| References replaced | No | Yes | No |
| Tables by LLM or Manual | LLM | Manual | Manual |
| **Total score excluding bibliography (%)** | **26%** | **19%** | **14%** |
| #1 Source (%) | frontiersin.org (5%) | frontiersin.org (4%) | frontiersin.org (2%) |
| #2 Source (%) | medrxiv.org (2%) | medrxiv.org (1%) | crossref source (1%) |
| #3 Source (%) | ncbi.nlm.nih.gov (2%) | ncbi.nlm.nih.gov (1%) | mdpi.com (1%) |
| #4 Source (%) | crossref source (1%) | crossref source (1%) | ncbi.nlm.nih.gov (1%) |
| #5 Source (%) | multicentredatabase.net (1%) | researchgate.net (1%) | researchgate.net (1%) |
| **Total score including bibliography (%)** | **37%** | **21%** | **25%** |

LE8: Life's Essential 8, BRI: Body Roundness Index

The LLM-rewritten manuscripts are presented in Supplementary Materials with watermarking and disclaimers. The watermarks and disclaimers were not included in the versions uploaded to iThenticate.

## Discussion

Plagiarism and related misconduct such as submissions of redundant manuscripts have been a constant in scientific publishing, and many checks and balances exist to guard against these problems. ^24,25^ The results presented here, however, are suggestive that these checks are not adequate in the GenAI era, given the sharp acceleration since 2022 in the number of redundant publications (2024 exhibiting a 17-fold change versus 2022, for example). Our workflow identified 411 total redundant publications from a single data source, NHANES, but it should be noted that the matching procedure was heavily automated, and better-concealed duplications would have escaped this process (a full manual text check of 15 000+ manuscripts being beyond the scope of this work). This has happened at the same time as wider growth in submissions putting editorial and peer reviewing processes under increased strain, ^26,27^ possibly increasing the dependence on automated systems such as iThenticate. The workflow also identified some journals as accepting a disproportionate number of redundant publications, which may reflect paper mills targeting journals where fabricated manuscripts have already been accepted, ^28^ perhaps reflecting perceived weaknesses in their editorial checks.

Our findings on duplicates illustrate the scale of the problem, but do not provide the mechanism by which it is happening. Here, we have demonstrated that ‘new’ syntactically different manuscripts can easily be generated that avoid text recycling or textual plagiarism. Whilst our work does not represent a systematic investigation of how easily plagiarism checks can be avoided, the three pairings chosen here did not exceed warning thresholds for individual source similarity scores of 5%, and were also free of ‘tortured phrases’, another indicator often used to detect problematic publications. ^29^ Interestingly, for all three synthetic manuscripts the Frontiers family of journals provided the highest ‘individual source similarity’ scores, which likely reflects Frontiers accepting more of these types of publications than other journals (Frontiers accepted 29% of the redundant publications identified in this work). Our synthetic manuscripts also passed thresholds despite our selecting highly duplicated pairings of exposures and outcomes, and the substantial time gap between publication and our submitting of the manuscripts to iThenticate.

Redundant publications can also amplify bad research. For example, vitamin D intake can be correlated with depression, but using NHANES data the association between insufficient serum vitamin D and depression has been shown by Diaz-Amaya et al. to be fully attenuated after adjusting for food security and diet quality. ^30^ Nonetheless, here we have identified three publications taking a simplistic single-factor approach reporting that serum vitamin D is associated with depression, findings which we regard as unsound. Therefore, misleading publications on serum vitamin D using NHANES outnumber the more thorough analysis in a 3:1 ratio. A simple comparison by Google Scholar shows citations for these publications in a 30:2 ratio. This is likely to be a major problem for those that are not domain experts – epistemic trespassers are much more likely to take the ‘weight’ of publication or citation counts as indicative of evidence. In so doing, epistemic trespassers can then further propagate low-quality results.

These findings extend previous work highlighting mass-manufactured low-quality research generated from publicly accessible datasets, leading to the literature being oversaturated with repetitive and potentially misleading findings. ^15,31^ Combined with the strong incentive to publish frequently (culturally for authors, and financially for journals), this has created a system where quantity is able to displace quality. There is growing evidence that these features have been exploited by “paper mills”, organisations that will produce manuscripts on a large scale, typically with minimal originality, and offer authorship slots to academic customers to meet the demand for rapid publication. ^32,33^ NHANES is an attractive target for these organisations: it is publicly available, has no registration or reporting requirements, and can be mined repeatedly with surface-level variations in study design. It is, however, important to recognise that the methods described in this work will not just be applicable to NHANES, albeit they may be easier to identify for this dataset due to the sheer volume of manuscripts. There is no reason why redundant submissions would not be enabled by GenAI for other health datasets, or for other types of research, such as systematic review papers, for example. We see risks that paper mills may migrate into these areas if negative attention around data resources such as NHANES limits publication opportunities. Epistemic trespassing is also likely to see growth, as researchers new to an area familiarise themselves with a topic using GenAI and see that it offers an easier route to publication.

Addressing these challenges will not be straightforward. COPE definitions do not fully capture this type of misbehaviour, as their categorisation of ‘redundant publication’ deals with the same authors submitting to multiple journals. ^8^ We contend that this definition does not reflect the reality that the true author is concealed by contract cheating, whereby the disclosed authors appear to be different (but have in fact purchased authorship from the same source). Proving this in individual cases is not easy, but we see no other plausible explanation for the aggregated surge in redundant publications described here.

### Limitations

Whilst the overall trends are informative and illustrate a growing problem, a number of limitations should be stressed. First, from reviewing manuscripts alone, it is not possible to differentiate between researchers with good intentions that have simply written papers where they lack the domain expertise to be aware of competitor groups and publications (epistemic trespassers), versus unethical actors such as paper mills deliberately using strategies such as simultaneous submission to multiple venues. Second, here we took a conservative approach to minimise false positives, by excluding associative studies that analysed specific subsets of data. This may understate the scale of the problem. Third, this work has identified the potential for using LLMs to quickly rewrite manuscripts. Here, however, we have shown illustrative examples. This is not a comprehensive analysis across a large number of manuscripts to identify how often tools such as iThenticate can be ‘fooled’. The evidence from the overall publication of duplicated topics, however, suggests that it is unlikely that iThenticate and similar tools are well prepared for the changing landscape, and it is the nature of LLMs that should a submission fail, the LLM can simply reword the document again until it passes. Fourth, our workflow was designed to test whether documents rewritten by LLMs would pass automated checks. We do not suggest that our documents would pass the desk editor or peer review checks in place at responsible journals, and further interventions and editing would be required to do so. Fifth, interventions were required to correct simple errors introduced, and this typically required an additional two hours of manual checking. The prompts used here were not able to produce new error-free manuscripts in a single click, albeit the reduction in time would in our view represent a massive time saving.

### Conclusion

These results demonstrate that current checks for redundant publications and plagiarism are no longer fit for purpose in the GenAI era. There are, however, few easy solutions. We expect greater emphasis on AI and other forms of detection from publishers as another ‘automated’ check, especially as publication volumes appear to be expanding beyond the ability of manual checks and balances such as peer review to cope. For known ‘templates’ such as NHANES, FAERS or two-sample Mendelian randomization (e.g. derived from FinnGen or the UK Biobank), we believe publishers need to be especially vigilant for formulaic manuscripts. In the long-run, greater co-operation between publishers for a register of submissions in these areas might help to reduce redundancy. We believe that updated / modified guidelines will be required from COPE to address these new innovations in paper mill production, especially to move away from current narrow definitions of redundant publication that focus on visible author overlap, for example. Without action to address these new GenAI related challenges, however, we expect the problem to continue to grow, making scientometric and meta-analyses more challenging, wasting publisher and peer reviewer resources, and potentially reducing the integrity of the scientific record.

## Supporting information

Supplementary Materials: Synthetic Manuscripts

## Supplementary Materials

The list of exposures and outcomes used to identify redundant publications is included in Supplementary Material, Table S1. The complete dataset of exposure versus outcome pairings including PMIDs is included in Supplementary Material, Table S2. All three synthetic manuscripts are additionally included as Supplementary Materials.

## Author Contributions’

Conceptualization, M.S. and A.B.; methodology, M.S. and A.B.; software, M.S.; T.S.; formal analysis, M.S. T.S. and D.M.; investigation, T.S. and D.M.; resources, M.S. and A.B.; data curation, T.S. and D.M.; writing—original draft preparation, M.S. T.S. and D.M.; writing— review and editing, M.S. and A.B.; visualization, M.S.; supervision and project administration, M.S. and A.B.; funding acquisition, M.S. All authors have read and agreed to the published version of the manuscript.

## Data Availability Statement

The data underpinning this work can be reproduced from the PubMed database at https://pubmed.ncbi.nlm.nih.gov/. The complete dataset of exposure versus outcome pairings including PMIDs for the 411 redundant publications included in this work is included in Supplementary Material, Table S2.

## Acknowledgements

The authors wish to acknowledge the work of the United2Act network in its work to highlight the threats to scientific integrity from paper mill activity. ^34^ The authors also wish to acknowledge the Collection of Open Science Integrity Guides (COSIG). ^35^ The authors also wish to declare that GenAI (Large Language Models) were used to prepare the synthetic manuscripts in this work, as described under Methods.

